# Outcomes and factors influencing care decisions in life-threatening fetal and neonatal anomalies

**DOI:** 10.1101/2020.07.12.20152280

**Authors:** Esther J. Lee, Simone Stenekes, Michael Harlos

**Affiliations:** Department of Pediatrics, University of British Columbia, University of Manitoba; Canadian Virtual Hospice, University of Manitoba; Pediatric Symptom Management & Palliative Service, Palliative Care Program, Winnipeg Regional Health Authority, University of Manitoba; Department of Family Medicine, University of Manitoba; Canuck Place Children’s Hospice, Vancouver, Canada

**Keywords:** palliative care, hospice, perinatal, end-of-life care, decision-making, lethal life-threatening anomalies, congenital anomalies, selective abortion

## Abstract

**Objective:** A retrospective chart review was undertaken to describe the outcomes following the diagnosis of a life-threatening fetal and neonatal anomaly.

**Study Design:** Criteria for a life-threatening anomaly included genetic conditions, renal and pulmonary diseases, central nervous system anomalies (CNS), and cardiac defects. Information that could impact decision making was collected from provincial databases

**Results:** 176 patients met the inclusion criteria. The majority of the diagnoses were in the genetic and CNS categories. 58% (n=103) decided to terminate the pregnancy. When a live birth occurred (n=39), the mean length of survival was 13.5 days (range 1-156 days), with one death occurring at home.

**Conclusions:** Ethnicity and geographical factors were associated with the decision to terminate a pregnancy. The involvement of the palliative care service was associated with decreased interventions. Further research to determine the needs of families would be helpful in identifying program priorities in perinatal palliative care.

## Introduction

With advancements in technology, it is possible to diagnose life-threatening anomalies in the prenatal period. This has had a profound impact on the practice of palliative care in the perinatal period (1, 2). In a 2002 multicenter review of pediatric patients receiving palliative care in Canada, 22% of the children had conditions arising in the perinatal period(3). Current statements advocating for pediatric palliative care focus on neonates and children, but not fetuses(4, 5). Although there are many overviews of perinatal palliative care and its importance, there is a paucity of perinatal palliative care research and little has been described regarding the decision-making factors of the parents facing the choice of termination(6-12). In 2007, Breeze and colleagues provided a description of palliative care for prenatally diagnosed life-threatening abnormalities (13). This prospective study of 20 patients in the United Kingdom was limited by small numbers and did not explore factors associated with decision-making. Calhoun and colleagues also provided an overview of a perinatal hospice program that cared for families whose fetus was diagnosed with a life-threatening congenital anomaly (14). This was also a small descriptive study undertaken within the United States. However, the population described in this study is different than in Manitoba, a province with a population of 1.2 million, with considerable culture diversity including those of First Nations and Hutterite heritage.

In order to gain knowledge about the perinatal palliative care population, a retrospective chart review was undertaken to provide a description of outcomes following the diagnosis of a life-threatening anomaly in the perinatal period. The main objective of this research was to determine the number and type of life-threatening fetal and neonatal anomalies diagnosed in the Canadian province of Manitoba within a five-year period (2005 – 2010). In addition, we evaluated the outcomes and potential factors influencing parental decision-making and also examined the involvement of pediatric palliative care services and its potential impact on treatment decisions.

## Methods

Ethical approval was obtained from the University of Manitoba Health Research Ethics Board. Site access and further approval was obtained through both hospitals where data collection took place.

Following a detailed literature search and discussion with the genetics and cardiology teams, it was determined which life-threatening anomalies would be included in the study (Table 1). Inclusion criteria for this retrospective chart review were pregnant women whose fetus had been diagnosed with a life-threatening anomaly, and neonates diagnosed with a life-threatening anomaly.

Several provincial databases were accessed in order to obtain information about patients meeting the inclusion criteria. These included the databases of medical genetics, fetal assessment, cardiology and pediatric palliative care. The health records databases of the two tertiary hospitals within the province were searched using diagnoses and ICD codes.

We collected demographic data (maternal age, urban or rural residence, cultural/spiritual background), subspecialty and allied health involvement, pregnancy and delivery details, and post-partum/postnatal and death details. The data were analyzed using SPSS.

## Results

### A. Demographics

A total of 176 patients with life-threatening anomalies were included in this chart review. A breakdown by diagnoses is presented in Table 2. The majority of the life-threatening diagnoses were in the genetic and CNS categories with Trisomy 18 being the most predominant. Ninety-five percent (n=167) of life-threatening anomalies were diagnosed prenatally.

### B. Outcomes

Fifty-eight percent of the pregnancies were terminated (n=103), while 41% were continued (n=71). For those who decided to continue the pregnancy and had a live birth (n=39), the mean length of survival was 13.5 days with a range of 1-156 days. Location of death included the labour and delivery unit (n=18), neonatal intensive unit (n=11), a rural hospital (n=5), inpatient unit (n=3), newborn nursery (n=2), postpartum unit (n=1), and home (n=1).

### C. Factors impacting decision making

Table 3 shows the factors that were associated with the decision to terminate. The decision to terminate was not associated with the diagnosis or the involvement of a particular subspecialty. Terminations occurred more often when the mother was of European or Asian origin (p=0.007) or if the father was of Asian origin (p=0.02). Terminations occurred less often if the father was of Hutterite or Mennonite origin (p=0.02). The stated faith of parents did not seem to influence the decision that was made (p=0.43 for mothers, and p=0.44 for fathers)

The mean age of the mothers at the time of initial assessment was 28 years. Maternal age was not associated with the decision to terminate or continue the pregnancy (p=0.27). The mean gestational age (GA) at diagnosis was 19.7 weeks, while the mean GA at diagnosis of those who decided to terminate was 18.7 weeks and those who decided to carry to term was 22.0 weeks (p=0.001). Other factors that were reviewed such as diagnostic methods (ultrasound, amniocentesis, chorionic villus sampling, echocardiogram, x-ray, MRI) were not associated with the decision to terminate or continue the pregnancy.

For those with documentation in their chart regarding reasons for their decision (n=37), the main reasons provided for their decision were: wishing comfort care (n=14); hopeless prognosis (n=10); carefully considering all options (n=8), and awaiting amniocentesis to confirm diagnosis (n=5).

### D. Pediatric palliative care (PPC) consultative service involvement

A total of 31 perinatal consultations occurred during the study period. Most of these consults occurred prenatally (65%, n=20), 13% (n=4) during labour, 16% (n=5) in the first week of life, and 6% (n=2) from the first week to the fourth week of life.

When compared with the patients in this study who did not have contact with pediatric palliative care, the involvement with PPC was associated with a decrease in the initiation of ventilatory support (p=0.025) and decreased length of stay in the NICU or nursery (p=0.019).

## Discussion

At the inception of this study, there was little published literature describing decision making for families faced with a diagnosis of a life-threatening fetal anomaly. This is the first Canadian study to provide a description of outcomes when a decision is made to continue the pregnancy.

The rate of termination of pregnancy in this study (58%) is similar to that reported by Breeze (60%). The mean gestational age (GA) at diagnosis was 19.7 weeks, which is similar to the study by Breeze (13) as well. However, this study found a difference of the mean GA between those who decided to terminate (18.7 weeks) and those who decided to carry to term (22 weeks, p=0.001). The reason for this disparity is not clear, and this has not been examined in other studies. One potential factor may be rural location, where timely access to early diagnostic ultasounds is not always available. The pregnancy termination policy in Manitoba at the time of this study was that termination was available in a pregnancy up to 19+6 weeks inclusive (15); after that date, each case is considered in its clinical context.

Our research is consistent with other work showing that the decision to terminate a pregnancy with a life-threatening anomaly was associated with specific ethnic backgrounds of the parents (16, 17). We also found that urban home location was associated with increased termination rates, which has not been reported in any previous studies. Although we included many factors that could impact decision making by parents, the majority of charts did not include detailed psychosocial information. Spiritual/religious background was only indicated in the charts of 25 mothers. Taking into account the minimal data obtained, there were not any statistically significant differences between various spiritual backgrounds (p=0.43). Unfortunately, the absence of data cannot be remedied in a retrospective chart study. Nevertheless, Howard suggests that declaring a specific faith background on a screening question does not delve into other personal values and pre-existing preferences, which could impact decision-making (18).

Recently there have been more studies evaluating the possible factors influencing decisions regarding pregnancies complicated by life-threatening fetal anomalies. A systematic review of the literature was undertaken by Jeon and colleagues regarding decisions to terminate after a prenatal diagnosis of a sex chromosome abnormality. From the review of 19 studies, they found that the factors common to both termination and continuation decisions are the specific type of sex chromosome abnormality, parents’ age (older women tended to continue the pregnancy while younger women tended to terminate), gestational age at diagnosis, providers’ expertise in genetic medicine and number of children/desire for (more) children. The factors that were associated with the decision to terminate were parents’ fear/anxiety and directive counseling. Factors associated with the decision to continue a pregnancy were parents’ socioeconomic status (low or middle depending on the study) and either Hispanic or Caucasian ethnicity (17). A group in Turkey found that the severity of the genetic abnormality and religion were important factors in decision making when there was a fetal chromosomal abnormality (n=38) (19). A group in France found that parental ethnicity (more terminations in families of French origin), gestational age at diagnosis (more terminations with earlier diagnoses), and the severity of the congenital heart disease were factors associated with parental decisions when there was fetal heart disease (n=188) (16). Asplin’s group in Sweden found that women decided to terminate a pregnancy based on the severity of the malformation. They also indicated that socioeconomic considerations and the doctor at the fetal care unit influenced the decision (20).

Other advances in perinatal palliative care have involved the development of a structured end-of-life curriculum for neonatal-perinatal postdoctoral fellows (21) and studying the various perceptions of health care workers regarding perinatal palliative care (22-24). Asplin’s group completed qualitative studies about the experience of women in whom a fetal malformation is detected by ultrasound examination. They found that the important factors for the women were timing, duration, and the manner of the intial dialog and ongoing support (25). The women also indicated that the improvements they wanted to see were in communicaton, in-depth understanding, compassion, complete follow-up routines and increased resources (26). Another important article is by Wilkinson discussing the importance of language and whether instead of the word “lethal”, other terminology like life-limiting or life-threatening should be used (27). The latter term is supported by the leading pediatric palliative care research group in Canada (28).

Our study showed an association between the involvement of palliative care and less aggressive interventions for the live births. Despite the small number of patients in this study this association supports the inclusion of palliative care as part of the interdisciplinary team and offering this service to all families with a fetus who has a life-threatening anomaly. Wool demonstrated that in various studies where parental response to perinatal palliative care was explored, the feedback was overwhelmingly positive (8). Bennett described in a case-study the importance of an palliative interdisciplinary approach to meet the complex bereavement needs of families (29).

It is important that all families be supported no matter what decision they make. A model program for perinatal palliative care services was detailed by a group in California but did not include how to support the family if they decided on termination or if the baby died prior to delivery (30). Wool performed a systematic review of parental outcomes after the diagnosis of a fetal anomaly and found that grief reaction was similar when termination was chosen compared to a spontaneous perinatal loss (8). Davies discovered that there were high levels of psychological stress in women who had terminations for a fetal anomaly and Kersting showed that post-traumatic stress and depressive symptoms were the highest in women who had a termination of a late pregnancy (31, 32). Perinatal palliative care has an opportunity to meet the needs of patients and families not just at the beginning of the diagnosis but also through bereavement. A recent study by Wool shows that most perinatal palliative care services in the United States have developed in the last 10 years (33). Although there is much to be done to enhance the care provided to families, the development of standards of care and ongoing research will hopefully improve the experience of families (34).

## Conclusion

Decision-making by parents regarding the approach to care for life-threatening fetal and neonatal anomalies is complex and multifactorial. This study shows an association between a decision to terminate a pregnancy and parental ethnicity as well as urban residency. We also found that involvement of the palliative care team was associated with fewer postnatal interventions and a shorter hospital stay. Prospective studies exploring the needs of families and caregivers from diagnosis through to bereavement are needed in order to determine the most appropriate interventions and resources supporting those facing life-threatening fetal and neonatal anomalies. Guidelines outlining care options and best practice for perinatal palliative care should be developed to ensure equitable access to supports for families and education for care providers.

## Data Availability

Data no longer available due to time since data collection/research completed

## Acknowledgements

The authors thank the Manitoba Institute of Child Health (MICH) and the Winnipeg Regional Health Authority Pediatric Palliative Care Grant for the financial support. We also acknowledge

- Dr. Bernie Chodirker for assistance with access to the genetics database and assisting with data interpretation
- Dr. Rasheda Rabbani at MICH for statistical support
- Stephanie Lelond, RN for undertaking part of the chart review

## Abbreviations

CI: confidence interval
GA: gestational age
ICD: international classification of diseases
IUD: intrauterine death
LR: likelihood ratio
PPC: pediatric palliative care

